# Global Catastrophic Risks and Capacity to Manufacture Key Pharmaceuticals: Case Study for a Highly Trade-Dependent Nation

**DOI:** 10.1101/2025.05.06.25327132

**Authors:** Nick Wilson, Peter Wood, Matt Boyd

## Abstract

**Introduction:** Human civilisation faces such global catastrophic risks as a: nuclear war, bioengineered pandemic, major solar storm, and volcanic winter. For some of these catastrophes, island nations may have relative survival potential but any collapse in international trade could also end critical imported goods such as pharmaceuticals. We aimed to explore the latter using the case study country of Aotearoa New Zealand (NZ).

**Methods:** We identified the 10 most extensively prescribed pharmaceuticals in NZ that are used for acute treatment (by annual prescription numbers). Based on modern synthesis pathways for these pharmaceuticals in the literature, we identified ingredients and then determined if these ingredients were currently produced in NZ.

**Results:** The results suggest that none of these 10 pharmaceuticals could be produced in NZ in a trade-ending catastrophe: paracetamol, omeprazole, amoxicillin, ibuprofen, aspirin, metoprolol succinate, salbutamol, prednisone, cetirizine hydrochloride, and amlodipine. This is primarily because NZ does not refine petrochemicals. For seven of these 10 pharmaceuticals the relevant catalysts or other specific chemical ingredients are also not mined or otherwise produced in NZ. There may, however, be some scope for the post-catastrophe scavenging of minerals for producing some catalysts.

**Conclusions:** This preliminary analysis suggests that none of the 10 most extensively prescribed pharmaceuticals used for acute treatments could be manufactured in this case study country after a trade-ending global catastrophe. To address this and other domains lacking in resiliency (eg, liquid fuel supply), a research programme for building shared resiliency with other neighbouring nations (eg, Australia) could be considered.

## INTRODUCTION

There is concern among experts about global catastrophic risk^1^ and the US Government recently requested a Report that has detailed the following threats: “artificial intelligence; asteroid and comet impacts; sudden and severe changes to Earth’s climate; nuclear war; severe pandemics, whether resulting from naturally occurring events or from synthetic biology; and supervolcanoes.”^2^ The risk of some of these may also be increasing eg, for pandemics.^2 3^

The risk of nuclear war may also have been increasing in recent years with repeated implied threats around nuclear weapons by the Russian leadership^4^ and deteriorating international relations between some of the nuclear weapon states (eg, the US and Russia; and the US and China). There is also the likely expiration of a key nuclear weapons treaty in 2026^5^ and a general lack of meaningful progress with nuclear disarmament.^6^ Furthermore, nuclear weapon states are typically either modernising and/or expanding their arsenals (eg, China^7^, North Korea,^8^ and Pakistan^9^).

Some island nations may be better placed than other nations to survive such catastrophes.^10 11 12^ But major challenges would likely include maintaining key aspects of modern society that depend on international trade (eg, food, liquid fuels, and industrial supplies), given that such trade could be severely disrupted or end entirely. In this particular study we look at just one such aspect: the supply of key pharmaceuticals.

In recent years, a number of high-income nations have recognised the vulnerability of their pharmaceutical supply chains. These concerns were particularly highlighted by the Covid-19 pandemic, with nations discovering high level dependencies on both imported active pharmaceutical ingredients (APIs) and finished medications. In response, the United States (US) in 2021 announced establishing public-private partnerships to identify and domestically manufacture 50-100 critical medications.^13^ Furthermore, the European Union (EU) has proposed a Critical Medicines Act to address severe shortages.^14^ Some countries within the EU have taken more concrete steps, with France implementing a “Pharmaceutical Sovereignty” initiative that includes restarting domestic paracetamol production (with production due to start in 2026).^15^ Similarly, in 2024 Sweden proposed state-run pharmaceutical manufacturing for essential medicines.^16^ Nevertheless, the dependency on international trade for APIs remains very high with Europe, for example, obtaining 60-80% of its APIs for generic medicines from China.^17^ For smaller countries (such as our proposed case study country of New Zealand), the economic barriers to comprehensive pharmaceutical independence are particularly substantial. Australia, for example, imports 90% of its medicines.^18^

### Background on the case study country: New Zealand

New Zealand is a high-income island nation where its government agencies have previously considered the threat of nuclear war,^19 20^ pandemics,^21^ and more recently severe space weather events that could damage the national electrical grid.^22^ But this country still remains extremely unprepared for such catastrophes as recent studies on nuclear war/winter indicate.^23 24^ Although in some global catastrophe scenarios it is plausible that some level of trade between New Zealand and Australia might continue – this is far from guaranteed given New Zealand’s complete dependence on imported liquid fuel for aircraft and cargo shipping.^23^ Furthermore, this country may have relatively little to offer Australia in terms of critical trade (that Australia doesn’t already produce), and it is also relatively far away from all other trading partners.

One of the major threats to health of New Zealanders from a global catastrophe could be in terms of food security – given the extreme dependency of industrial agriculture on imported liquid fuels.^24^ This is despite the apparent national self-sufficiency in food.^25^ New Zealand research has started to consider this threat in terms of estimating local biofuel production to keep agricultural machinery running,^24^ optimal crop selection,^26^ and optimal use of urban and peri-urban agriculture.^27^ But other threats to health from such global catastrophes have only been explored in relatively outdated work from the 1980s.^19 20 28^ In particular, the topic of pharmaceuticals has not been studied in detail, even though this is a likely area of high vulnerability since:

- The existing pharmaceutical industry in New Zealand does not currently produce major pharmaceuticals from source ingredients. Instead, it is focused on secondary manufacturing and formulation, packaging of imported active ingredients, and quality control and testing.
- The recent ending of oil refining in New Zealand^29^ (despite continuing to produce and export crude oil), has curtailed potential domestic production of petroleum-derived ingredients for pharmaceutical manufacture. There are also no industrial plants using coal-to-chemicals or coal-to-liquids technologies. Methanol is, however, manufactured from natural gas in Taranaki,^30^ and there is a wood pyrolysis plant in Timaru.^31^ But the latter only produces charcoal and not chemical by-products potentially relevant to pharmaceutical production (eg, phenolics and furans).
- Industrial production of other chemicals relevant to the pharmaceutical industry is also relatively limited. However, hydrogen gas is produced at various sites (eg, from a geothermal plant^32^), and sulphuric acid is produced as part of fertiliser production (superphosphate).^33^

## METHODS

The most extensively used pharmaceuticals in New Zealand by annual numbers of dispensed prescriptions,^34^ were selected. These were then categorised to produce a list of 10 that were used for acute treatment, as opposed to chronic disease management (Table 1). The focus on acute treatment may reflect potential lifesaving use in some cases, but also the production demands would typically be lower given that the volume of pharmaceuticals needed for short courses of acute treatment is typically much less than for chronic disease management. We did not consider the list of “essential medicines” in the World Health Organization’s list^35^ as this is very extensive (591 drugs and 103 therapeutic equivalents), and is not prioritised.

**Table 1:** Most extensively prescribed medicines in the 2022/2023 financial year in New Zealand^34^ used to identify the 10 most relevant medicines for treating acute conditions. (ranked by annual number of prescriptions dispensed according to the national Pharmaceutical Management Agency of the New Zealand Government, [Pharmac])

| Medicine | Therapeutic group (Pharmac) | Prescriptions (annual number) | Included in this study if relevant to acute treatment |
| --- | --- | --- | --- |
| Paracetamol | Analgesics | 3,460,000* | Yes – for acute pain relief; fever reduction (suitable for children) |
| Atorvastatin (statin) | Cardiovascular | 1,840,000 | No – since used for the *prevention* of cardiovascular disease |
| Omeprazole (proton-pump inhibitor for heart burn/acid reflux) | Alimentary | 1,690,000* | Yes – for acute gastritis and treating gastric ulcers |
| Amoxicillin (antibiotic) | Anti-infectives | 1,230,000 | Yes – for infections (potentially also lifesaving eg, severe bacterial pneumonia) |
| Ibuprofen (anti-inflammatory) | Analgesics | 1,200,000* | Yes – for acute pain relief |
| Colecalciferol (vitamin D) | Musculoskeletal | 1,110,000* | No – typically used for *prevention* of osteoporosis |
| Aspirin | Antithrombotics | 1,100,000* | Yes – for managing a heart attack or stroke (potentially lifesaving); also (in adults) acute pain relief and fever control. (Low dose aspirin is also used in non-acute disease management, ie, for the secondary prevention of cardiovascular disease.) |
| Metoprolol succinate (an antihypertensive) | Cardiovascular | 920,000 | Yes – for managing acute cardiac conditions such as arrhythmias and heart failure (potentially lifesaving); and managing angina. (It is also used in non-acute disease management, ie, for treating high blood pressure, a risk factor for cardiovascular disease.) |
| Salbutamol (a bronchodilator) | Respiratory | 860,000 | Yes – for acute asthma attacks (potentially lifesaving) |
| Levothyroxine (thyroid hormone) | Hormones | 710,000 | No – typically used for *managing* hypothyroidism. It is only lifesaving in extremely rare forms of hypothyroidism ie, myxedema coma. |
| Prednisone (steroid) | Hormones | 700,000 | Yes – for severe allergic reactions, autoimmune crises and asthma attacks (potentially lifesaving); acute management of chronic obstructive pulmonary disease, rheumatological conditions, and other diseases |
| Cetirizine hydrochloride (antihistamine) | Antihistamines | 690,000* | Yes – for allergic reactions; urticaria |
| Amlodipine (calcium channel blocker) | Cardiovascular | 680,000 | Yes – for certain types of angina. (It is also used in non-acute disease management, ie, for treating high blood pressure, a risk factor for cardiovascular disease.) |

\*Actual usage will probably be higher than suggested in this table as these particular pharmaceuticals are also sold over-the-counter in New Zealand eg, in pharmacies and/or supermarkets.

We then conducted Google Scholar searches in December 2024 to identify modern methods of synthesis and the associated ingredients for each of the 10 selected pharmaceuticals. These ingredients were then assessed in terms of New Zealand capacity to supply (eg, if particular minerals that are used for ingredients or catalysts are mined in New Zealand^36^).

### Ethics Statement

This study did not require ethical review as it was about the potential for pharmaceutical manufacturing in post-catastrophe situations and did not involve any human participants.

## RESULTS

Results for each of the 10 most extensively prescribed pharmaceuticals used for acute treatment in New Zealand are shown in Tables 2 and 3. These 10 covered the therapeutic groups (as classified by the national Pharmaceutical Management Agency of the New Zealand Government [Pharmac]), of: analgesics (n=2), alimentary (1), anti-infectives (1), antithrombotic (1), cardiovascular (2), respiratory (1), hormones (1) and antihistamines (1). Of note, however, is that some of these pharmaceuticals have roles in multiple groups (eg, aspirin also for analgesia and prednisone also for the respiratory group and for rheumatological conditions etc).

**Table 2:** Summary results relevant to potential New Zealand (NZ) production capacity for the 10 selected pharmaceuticals after a trade-ending global catastrophe (see Tables 1 and 3 for additional details)

| Medicine (ranked as per Table 1) | Modern synthesis requires products from petrochemical refining that does not occur in NZ* | Modern synthesis requires other chemicals from compounds not produced in NZ | Modern synthesis requires catalysts from minerals not mined in NZ** |
| --- | --- | --- | --- |
| Paracetamol | Yes | No | Yes (platinum) |
| Omeprazole | Yes | No | Yes (molybdenum) |
| Amoxicillin | Yes | No | No |
| Ibuprofen | Yes | Yes (fluorite) | Yes (nickel, bauxite, palladium) |
| Aspirin | Yes | No | No |
| Metoprolol succinate | Yes | No | No |
| Salbutamol | Yes | Yes (lithium, bauxite) | Yes (palladium) |
| Prednisone | Yes | Yes (bauxite, chromite) | Yes (nickel, bauxite) |
| Cetirizine hydrochloride | Yes | Yes (borax) | No |
| Amlodipine | Yes | No | Yes (palladium) |

\* Excluding the production of methanol which does occur in New Zealand.
\*\* However, there could be scope for some minerals to be scavenged in a post-catastrophe environment, eg, platinum and palladium from vehicle catalytic converters.

**Table 3:**
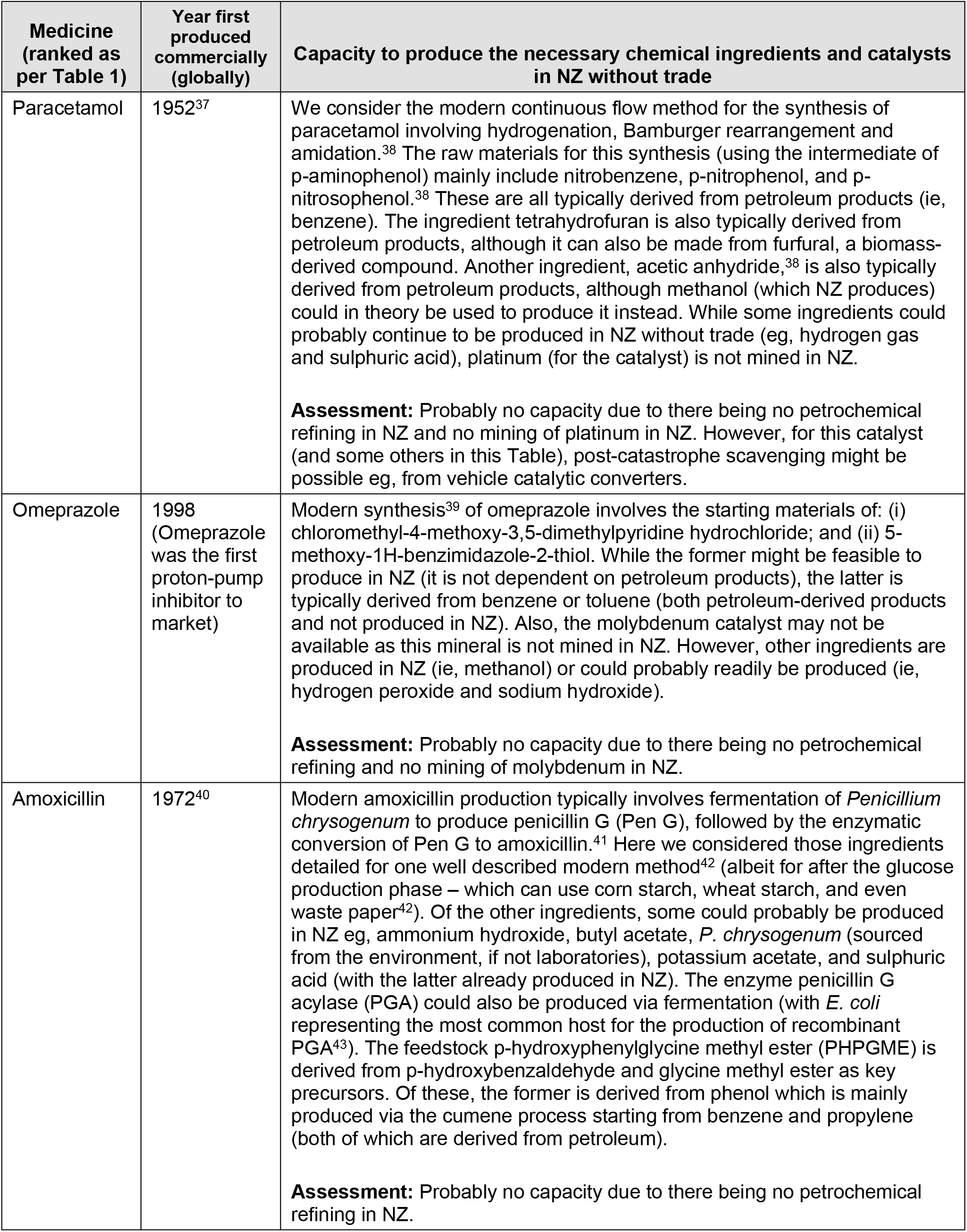
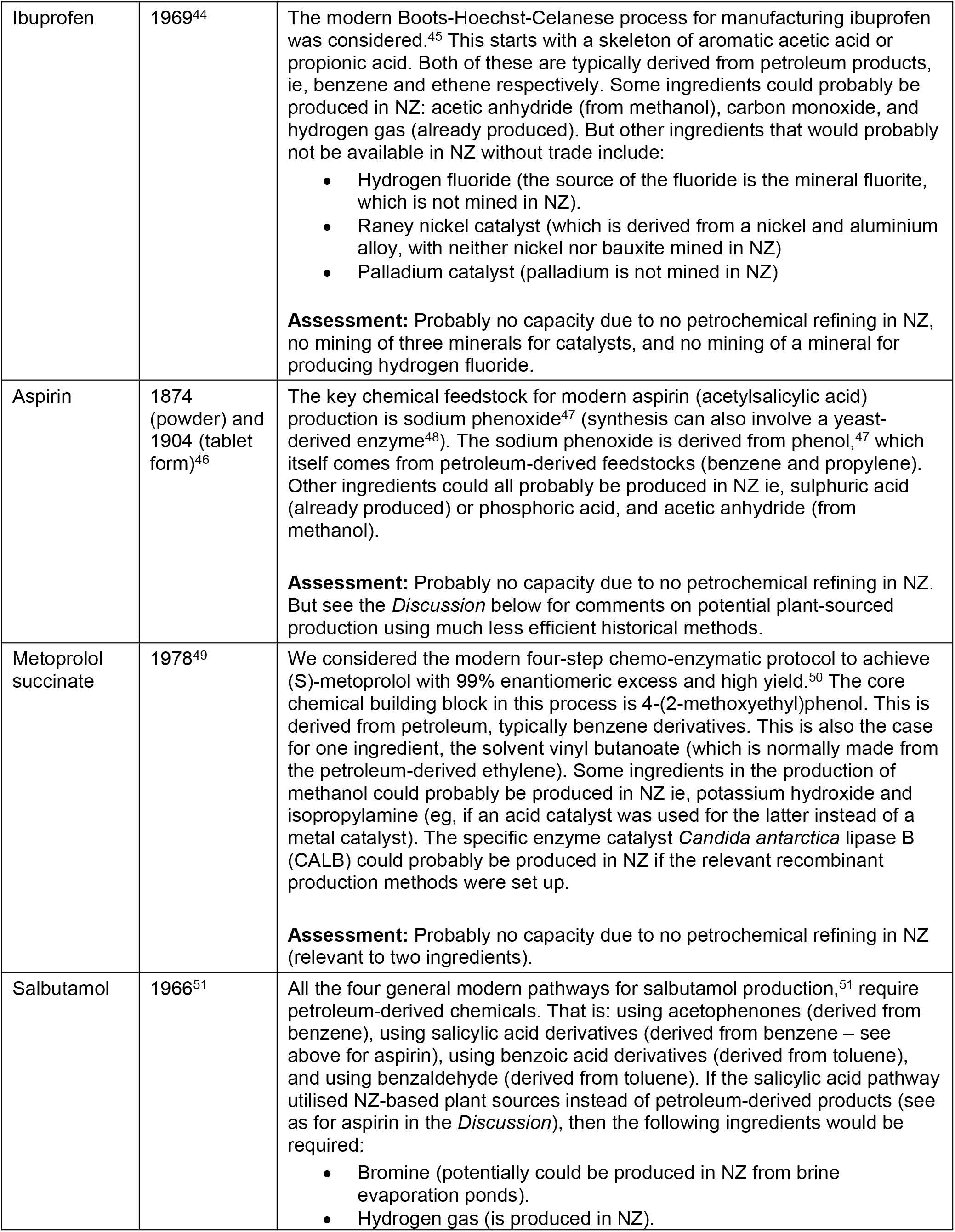
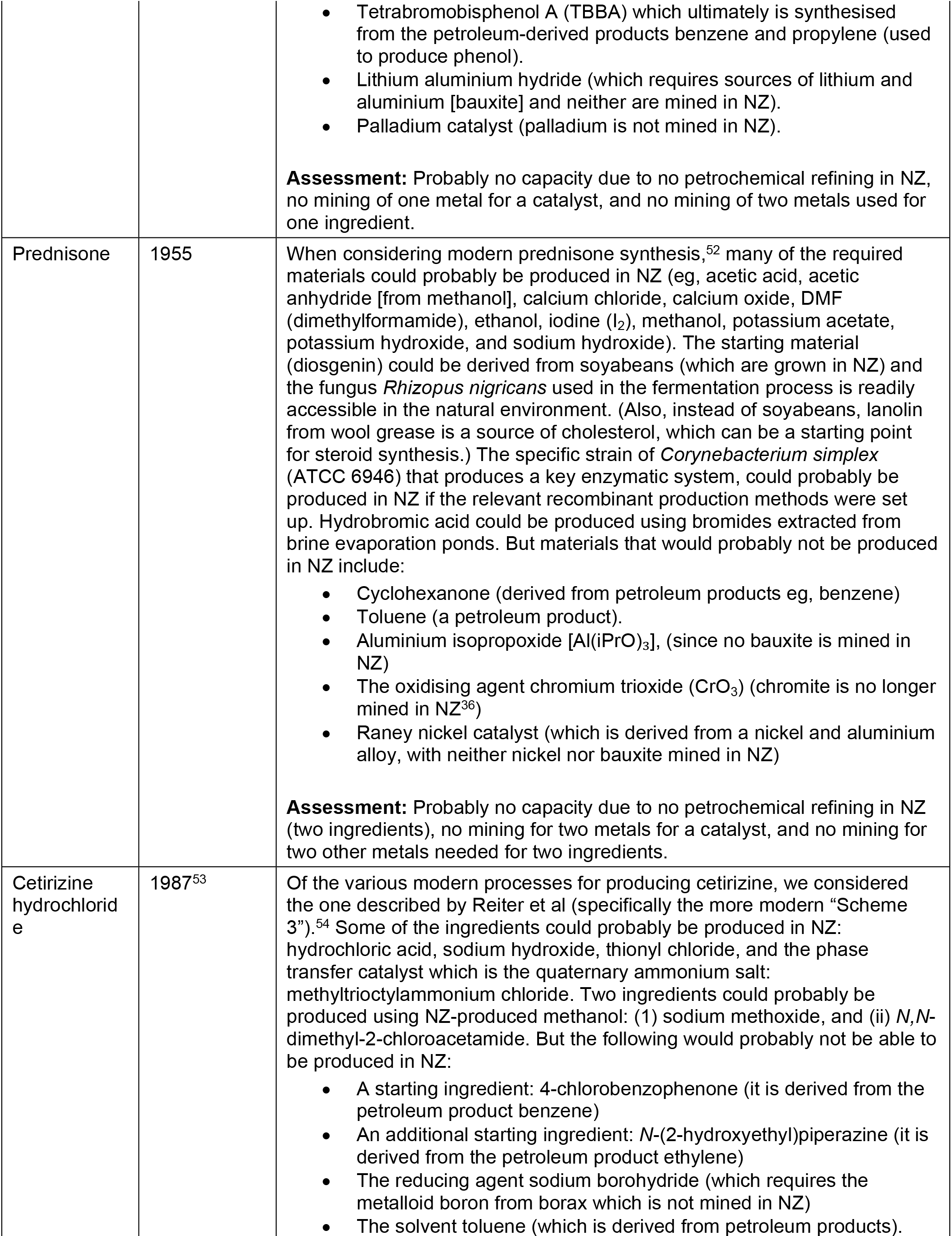
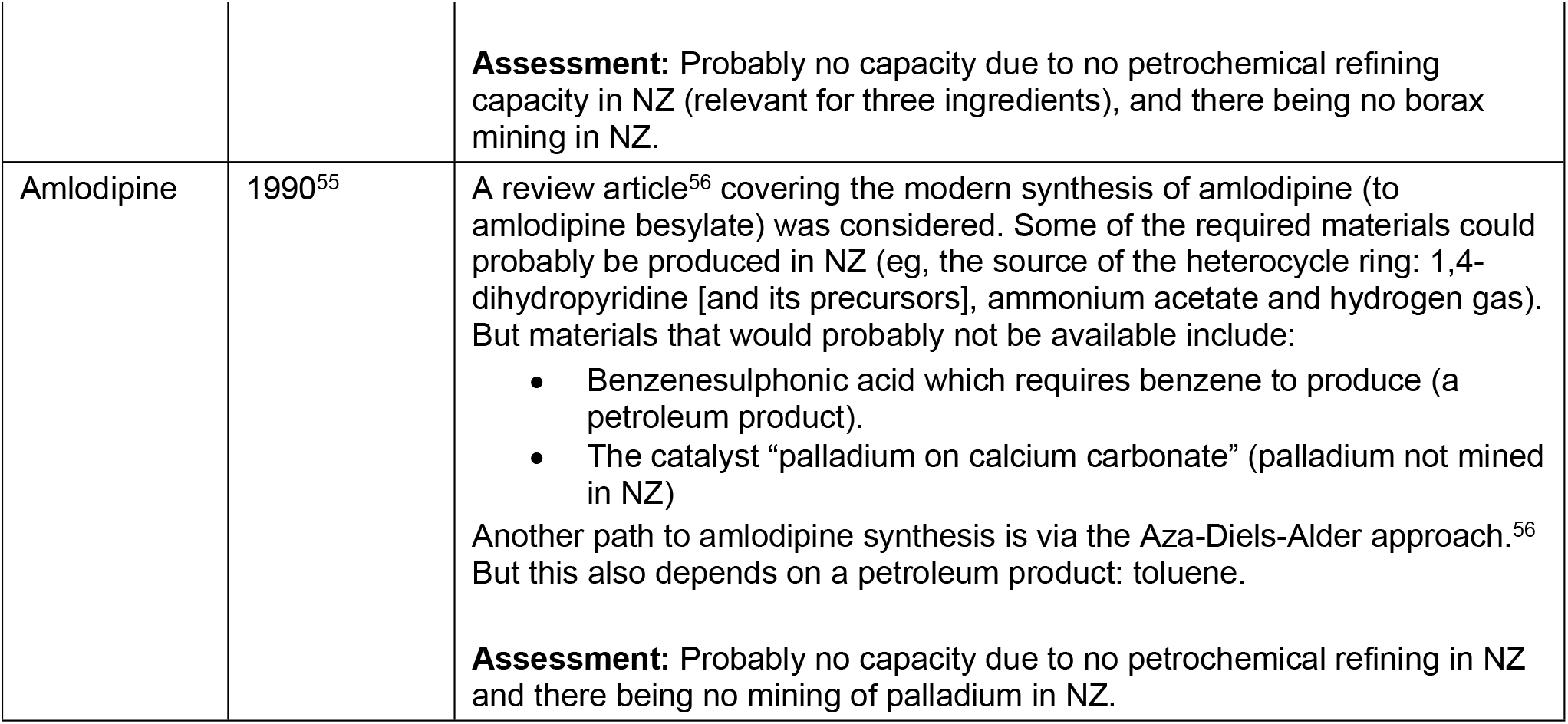
More detailed results relevant to potential New Zealand production capacity for the 10 selected pharmaceuticals in a trade-ending global catastrophe situation (see Table 1 for additional details)

The median year at which these pharmaceuticals were first available in the market anywhere in the world was 1971 (Table 3). But the range for these years was wide, at 1874 for aspirin in powder form, and 1998 for omeprazole.

A summary of the results in Table 2 suggests that none of these 10 pharmaceuticals could probably be manufactured using modern synthesis methods after a trade-ending catastrophe since New Zealand has no petrochemical refining capacity (other than methanol production). In addition, the modern synthesis of seven of the 10 pharmaceuticals would also probably not be possible due to catalysts or other specific chemical ingredients not being mined or otherwise produced in New Zealand (Table 2). Nevertheless, some of the catalysts may only be required in small amounts and so could potentially be scavenged in the post-catastrophe period within New Zealand eg, from vehicle catalytic converters.

## DISCUSSION

### Main findings and interpretation

This study suggests that after a trade-ending catastrophe, none of these 10 pharmaceuticals used for acute treatment could be manufactured using modern synthesis methods in New Zealand. This is primarily because this country has no petrochemical refining capacity, but also because it does not produce all the necessary catalysts or other specific chemical ingredients. Therefore, after such catastrophes, and once imported pharmaceutical stocks had run out, there would probably be increased deaths from infections, heart disease, stroke, and asthma; as well as increased morbidity (eg, from pain and prolonging the relevant illnesses detailed in Table 1). In terms of the health loss from untreated infectious disease, this burden would probably fall particularly on Māori (indigenous), Pacific peoples and socioeconomically deprived New Zealanders (who already have much higher current relative burdens^57^). Such inequities also exist for Māori and Pacific peoples for cardiovascular disease burdens.^58^

If modern synthesis methods were not used, could some of these pharmaceuticals still be produced in New Zealand? One possibility is aspirin given that alternative (non-petroleum) sources for salicylic acid include plants containing salicin eg, willow and meadowsweet,^46^ with the former commonly found in New Zealand. Aspirin manufacture is relatively simple once salicylic acid is available (at about high school student level).^59^ But if manufacturing involved plant-sourced salicylic acid, it would probably be far less efficient and more expensive than modern day synthesis methods. For example, there was a drop to a “tenth of the price” with industrial synthesis in 1874, relative to when extraction from willow was used.^46^

### Study strengths and limitations

Although preliminary in nature, a strength of this study is that it is the first to look in any detail at the issue of pharmaceutical production after trade-ending global catastrophes (to our knowledge). Nevertheless, the following limitations apply:

- The 10 selected pharmaceuticals were based on the number of prescriptions dispensed for medicines that had roles in treating acute conditions. But this was simplistic given that: (i) prescriptions numbers are only a crude indicator of volume and also do not account for non-adherence and other wastage; (ii) some of the selected pharmaceuticals are also used for non-acute treatments (eg, aspirin, metoprolol and amlodipine – see Table 1); and (iii) some of them can be purchased over-the-counter (eg, six of those in Table 1) and so are not counted in the “prescription data” that we considered. There was also no quantified consideration of life-saving potential as opposed to symptom relief for non-fatal conditions.
- As discussed for aspirin, this study focused on modern synthesis methods for these pharmaceuticals and did not explore alternative less efficient methods and historic methods. In particular, although some catalysts can be considered indispensable, substitute catalysts for many chemical reactions are often possible with trade-offs in terms of reaction efficiency.^60^ Also of note is that artificial intelligence is assisting with chemical substitution,^61^ as are “green chemistry” developments.^62^
- Options for the post-catastrophe scavenging of minerals not mined in New Zealand, were not fully explored, and yet this might be feasible if only small amounts of catalysts are required. For example, platinum and palladium could potentially be scavenged from vehicle catalytic converters (and platinum also from electrical equipment and jewellery). Nickel could be scavenged from stainless steel items and various industrial equipment. Also, where relevant minerals had previous been mined in New Zealand (eg, chromite^36^), mining operations could potentially be restarted if some ores still existed at recoverable levels. There may also be stockpiles of imported bauxite available if these were diverted from those held by the Tiwai Point aluminium smelter in Southland (relevant for ibuprofen, prednisone and salbutamol production). Similarly, stockpiles of imported fertiliser could be used as a source of boron (for cetirizine production) and stockpiles of imported fluoride, used for water fluoridation, could be used to produce hydrogen fluoride (for ibuprofen production). Even then, the capacity to turn such minerals and chemicals into usable ingredients in pharmaceutical manufacture would depend on the available expertise, refining capacity and manufacturing capacity.
- This analysis did not consider alternative treatments to modern pharmaceuticals. For example, rongoā Māori (traditional healing system) provides a range of alternative treatments based on rākau (plants).^63^ Also, in the European tradition, an example is the use of “medicated cigarettes” containing plants in the nightshade family (Solanaceae) for treating asthma, as used in the 19^th^ and 20^th^ centuries.^64^ Indeed, a review published in 2013 considered the effectiveness of *Datura stramonium* in treating asthma.^65^ However, such plants have toxicity risks,^65^ and misuse potential,^66 67^ and so might need medical supervision with administration.

### Potential further research and policy responses

In this unfunded research we took a fairly simple approach to pharmaceutical selection that involved prescription numbers for acute treatments. If there was government-funded research in the future, then a more sophisticated approach could prioritise a larger number of commonly prescribed pharmaceuticals (along with anaesthetics etc). Possible prioritisation could be informed by modelling the annual deaths prevented or annual quality-adjusted life years (QALYs) saved by different pharmaceutical treatments. If input from a citizen’s panel/assembly was included in the prioritisation process, then it could start to capture societal values eg, perhaps prioritising the production of antibiotics for saving the lives of essential workers over production of statins to manage risk factors such as elevated blood lipids.

All the above limitations with this preliminary analysis would suggest the need for more in-depth research. One of the more promising options might be for the New Zealand and Australian Governments to jointly plan for shared post-catastrophe production of key pharmaceuticals and the ability to trade them between each other by ship or aircraft. This trans-Tasman approach has already been suggested in terms of mRNA vaccine development for responding to future pandemics.^68^

Australia is much better positioned than New Zealand for pharmaceutical production since it still has oil refining capacity and its chemical industry and pharmaceutical industries are larger. Australia is even the source of 37% of the world’s licit morphine supply with opium poppy farms in Tasmania.^69^ The New Zealand Government could contribute funding for any Australia-based preparations and potentially provide some ingredients, but it could also focus on ensuring the viability of post-catastrophe trans-Tasman trade. For example, New Zealand-produced biofuels (eg, from canola cropping)^24^ could be used to keep cargo ships functioning in the absence of imported liquid fuels.

If the Australian Government was not interested in such joint planning, New Zealand Government commissioned research could still explore options for relevant plans for just New Zealand. This could especially focus on domains where some production may be more feasible (eg, aspirin production from plant-based sources) or adapting existing industrial infrastructure to produce the relevant ingredients. Examples might include modifying the current wood pyrolysis plant in Timaru to produce phenols and furans; or modifying the Glenbrook steel plant to produce benzene/phenol from coke gas. Even more expensive options would be building a micro-refinery for oil or coal tar.

### Conclusion

This preliminary analysis found that none of these 10 extensively used pharmaceuticals could probably be produced using modern synthesis methods in this case study country (New Zealand) after a trade-ending catastrophe. This is primarily because the country does not refine petrochemicals. To address this and other domains lacking in resiliency (eg, liquid fuel supply), a research programme for building shared resiliency with other neighbouring nations (eg, Australia) could be considered.

## Data Availability

All relevant data are within the manuscript and its Supporting Information files.

## Acknowledgements

Nil.

